# Then and NOW: A Prospective Population-Level Validation of the Abbott ID NOW SARS-CoV-2 Device Implemented in Multiple Settings for Testing Asymptomatic and Symptomatic Individuals

**DOI:** 10.1101/2022.04.30.22274189

**Authors:** William Stokes, Allison A. Venner, Emily Buss, Graham Tipples, Byron M. Berenger

## Abstract

**IMPORTANCE:** Since December, 2020, the ID NOW was implemented for use in 4 different populations across Alberta: in mobile units as part of community outbreak response, COVID-19 community collection sites, emergency shelters and addiction treatment facilities (ES), and hospitals.

**OBJECTIVE:** Diagnostic evaluation of the ID NOW in various real world settings among symptomatic and asymptomatic individuals

**DESIGN:** Depending on the implemented site, the ID NOW was tested on patients with symptoms suggestive of COVID-19, asymptomatic close contacts or asymptomatic individuals as part of outbreak point prevalence screening. From Jan – April, a select number of sites also switched from using oropharyngeal swabs to combined oropharyngeal + nasal (O+N) swabs. For every individual tested, two swabs were collected: one for ID NOW testing and the other for either reverse-transcriptase polymerase chain reaction (RT-PCR) confirmation of negative ID NOW results or for variant testing of positive ID NOW results.

**RESULTS:** A total of 129,112 paired samples were analyzed (16,061 RT-PCR positive). 81,697 samples were from 42 COVID-19 community collection sites, 16,924 from 69 rural hospitals, 1,927 from 9 ES, 23,802 samples from 6 mobile units that responded to 356 community outbreaks, and 4,762 from 3 community collection sites and 1 ES using O+N swabs for ID NOW testing. ID NOW sensitivity was highest among symptomatic individuals presenting to community collection sites [92.5%, 95% confidence interval (CI) 92.0-93.0%, n=10,633 RT-PCR positive] and lowest for asymptomatic individuals associated with community outbreaks (73.9%, 95% CI 69.8-77.7%, n=494 RT-PCR positive). Specificity was greater than 99% in all populations tested, but positive predictive value (PPV) was lower among asymptomatic populations (82.4–91.3%) compared to symptomatic populations (96.0-96.9%). There was no statistically significant differences in sensitivity with respect to age, gender, NP vs OP swab for RT-PCR confirmation, variants of concern, or with combined oropharyngeal and nasal swabs using COVID-19 ID NOW testing.

**CONCLUSIONS:** Sensitivity of ID NOW SARS-CoV-2 testing is highest when used on symptomatic community populations not seeking medical care. Sensitivity and PPV drops by approximately 10% when tested on asymptomatic populations. Using combined oropharyngeal and nasal swabs did not improve ID NOW performance.

## INTRODUCTION

The ID NOW (Abbott, Illinois, United States) is approved by the United States Food and Drug Administration (FDA) Emergency Use Authorization for the point of care, rapid detection of severe acute respiratory syndrome-coronavirus-2 (SARS-CoV-2) in individuals who are within the first 7 days of symptom onset.^1^ The ID NOW assay uses isothermal nucleic acid amplification of a region of the viral RNA-dependent RNA polymerase (RdRp) to detect the presence of SARS-CoV-2, with results available in under 15 minutes. Clinical specimens approved by U.S. FDA for testing include nasal, oropharyngeal, and nasopharyngeal swabs (NP), which must be tested on the Abbott ID NOW either immediately or within one hour of collection. Specimens placed in viral/universal transport media (UTM) are not valid for testing by the Abbott ID NOW.^1^

There are many studies that have evaluated the Abbott ID NOW. Its analytical sensitivity approximates 250-500 copies/mL and is significantly higher compared to other lab based RT-PCR platforms, which are typically under 200 copies/mL.^2,3^ Pooled clinical sensitivity of the ID NOW from 13 studies, compared to RT-PCR, was 73.0% (95% CI 66.8% - 78.4%), and specificity was 99.7% (95% CI 98.7% - 99.4%).^4^ However, these studies were limited in sample size and heterogeneous in design, such as in differences in specimen type and RT-PCR reference method used. There is also a paucity of data on ID NOW performance in asymptomatic individuals. This is important because ID NOW performance may be negatively affected among asymptomatic individuals with COVID-19, given the higher cycle threshold (Ct) values observed in this population.^5^

This study sought to accomplish two goals. The first was to prospectively evaluate the clinical performance of the ID NOW, compared to RT-PCR, after its implementation for SARS-CoV-2 testing in various settings across the province of Alberta, Canada (4.4 million people). Based on its rollout, we evaluated the clinical sensitivity and specificity of the ID NOW in these four settings:

1. Symptomatic individuals or asymptomatic close contacts presenting to community COVID-19 assessment/swab centres.
2. Symptomatic inpatients or Emergency Department patients.
3. Symptomatic individuals in emergency shelters and addiction treatment facilities.
4. Symptomatic or asymptomatic individuals associated with community outbreaks (majority in continuing care centres).

The second aim of this study was to compare the differences in sensitivity/specificity between the use of oropharyngeal swabs vs combined oropharyngeal and nasal (O+N) swabs for ID NOW testing in two different COVID-19 testing settings: assessment centres and emergency shelters and addiction treatment facilities. Although oropharyngeal swabs have slightly lower sensitivity compared to nasopharyngeal swab for SARS-CoV-2 detection by RT-PCR, several studies have demonstrated improved sensitivity with O+N swabs.^6-8^

## METHODS

Since December 4, 2020, the ID NOW was gradually implemented across Alberta in the following sites:

1. 42 COVID-19 Alberta Health Services (AHS) Public Health assessment/swabbing centres, located in all regions of Alberta: testing of symptomatic individuals and asymptomatic close contacts. These are the primary locations for community patients not needing medical attention to get tested for COVID-19 in Alberta. Testing and swabbing was performed by assessment centre staff (e.g. nurses) within Alberta Precision Laboratories (APL) approved Point of Care Testing (POCT) programs.
2. 69 rural hospitals located across Alberta: testing of symptomatic inpatients or ED patients. Swabbing was performed by physicians, nurses, or respiratory therapists. Testing was performed in a College of Physician and Surgeons of Alberta (CPSA) accredited hospital laboratory (APL).
3. 9 emergency/homeless shelters and addiction treatment facilities, located in urban centres in Alberta: testing of symptomatic residents. Testing and swabbing performed on site by registered nurses.
4. 6 mobile units that accessed all regions of Alberta: testing of symptomatic or asymptomatic residents or staff, all from organizations on outbreak (majority were continuing care sites). Swabbing and testing performed by mobile nursing team staff on-site in retrofitted vans.

Individuals were given the option to have POCT by ID NOW and routine testing, or routine testing alone. All individuals tested with the ID NOW had two parallel swabs collected. The first swab collected was either a NP swab or oropharyngeal (OP) swab, which was placed in UTM (Yocon Biology, Beijing, China or GDL Korea Co. Ltd, Seoul, Korea) for RT-PCR, and transported to an accredited laboratory at room temperature and stored at 4°C until processing. The second swab was an OP swab for ID NOW testing (using the OP swab provided in the ID NOW kits). The OP swab for ID NOW testing was always collected second to ensure all individuals had a sample available for RT-PCR (i.e. in case the individual refused the second NP or OP swab). If the ID NOW test was negative, the second swab was sent for confirmatory RT-PCR testing. If the ID NOW test was positive, the second swab was either sent for storage (if collected before February 1, 2021) or for variant of concern screening (VoC) testing. Samples sent for storage (−70°C) were tested at a later date for SARS-CoV-2 using RT-PCR. All RT-PCR tests sent for variant testing were done within approximately 72 hours from time of collection.

All RT-PCR testing was performed on the APL Public Health Laboratory E gene PCR or on a Health Canada and FDA approved commercial assay.^9^ Commercial assays were dependent on the testing lab and included the Allplex (Seegene, Seoul, South Korea), BDMax (Becton Dickinson, NJ, United States), Panther (Hologic, MA, United States), GeneXpert SARS-CoV-2 or SARS-CoV-2/Influenza/RSV (Cepheid, CA, United States), Cobas 6800 System (Roche Molecular Systems, CA, United States) and Simplexa (DiaSorin, Saluggia, Italy). All samples sent for VoC screening were tested with the ProvLab E gene assay to determine if sufficient viral load was present for VoC testing. E gene RT-PCR results from our lab-developed test were considered positive for SARS-CoV-2 when E gene cycle threshold (Ct) value was <35. If the Ct was ≥35, amplification from the same eluate was repeated in duplicate and was considered positive if at least 2/3 results had a Ct <41.

All personnel performing the ID NOW swabbing/testing were trained healthcare workers (HCW), who were previously trained in NP and oropharyngeal swab collection. At time of collection, they asked and recorded whether the patient had symptoms or was asymptomatic. All sites and HCW were trained on the ID NOW collection, transport, and testing processes, at least according to the manufacturer’s instructions, prior to ID NOW implementation. AHS staff were trained according to the APL POCT program, which meets CPSA accreditation standards. Each ID NOW device underwent a verification process, which included testing 3 positive ID NOW control swabs and 5 negative ID NOW control swabs on the ID NOW instrument before use. One positive control and one negative control swab were tested on the ID NOW instrument after each new box of ID NOW kits was opened, after each new HCW was trained on the instrument, and, after each instrument was transported to a different site.

All ID NOW samples with parallel RT-PCR results documented were included in our study. Results without proper documentation of testing location, or without either confirmatory RT-PCR or variant testing were excluded.

Between Jan 26 and April 12, 2021, 3 assessment centres and one shelter and addiction treatment facility switched from using oropharyngeal swabs to O+N swabs for ID NOW testing. The swab used remained the same (swab provided in Abbott ID NOW kits) and instructions for swabbing was provided to individual sites (see supplementary material for instruction details). All other protocols, as outlined above, remained in place.

Data was pulled from our provincial laboratory’s centralized electronic database containing SARS-CoV-2 results for all publicly funded testing in the province except for border testing. Sensitivity and specificity of the ID NOW was calculated with Clopper-Pearson 95% confidence intervals. Statistical analysis was performed using Pearson Chi-squared for categorical variables and t-test for continuous variables using STATA (version 14.1).

The University of Alberta Research Ethics board approved this study (Pro00111835).

## RESULTS

A total of 133,919 results were identified between December 4, 2020 and November 24, 2021. 4807 samples were excluded: 190 did not have testing location recorded, 375 did not have an ID NOW result recorded, 3 had invalid ID NOW results, 2,417 ID NOW results did not have parallel RT-PCR results recorded, and 1,822 positive ID NOW results did not have subsequent variant testing performed. The remaining 129,112 paired samples were analyzed.

### ID NOW testing using oropharyngeal swabs

124,350 samples (15,649 RT-PCR positive) were analyzed. 81,697 samples were from 42 assessment centres, 16,924 samples were collected from 69 rural hospitals, 1,927 samples were from 9 shelters and addiction treatment facilities, and 23,802 samples were from 6 mobile units that responded to 356 outbreaks. Baseline characteristics of these samples is provided in Table 1. Results, compared to RT-PCR, are provided in Table 2 and Figure 1. ID NOW sensitivity was highest among symptomatic individuals presenting to assessment centres [92.5%, 95% confidence interval (CI) 92.0-93.0%, n=10,633 RT-PCR positive], and lowest for asymptomatic individuals associated with community outbreaks (73.9%, 95% CI 69.8-77.7%, n=494 RT-PCR positive). There was an approximate 10% decrease in sensitivity when testing asymptomatic vs symptomatic populations. Specificity was greater than 99% in all populations tested.

**Table 1:** Characteristics between individuals tested with ID NOW SARS-CoV-2 using oropharyngeal swabs or combined oropharyngeal and nasal (O+N) swabs.

| Site | Symptoms | Mean age (median, range) | Male gender (%) | NP swab used for parallel RT-PCR testing* (%) |
| --- | --- | --- | --- | --- |
| **OROPHARYNGEAL SWAB FOR ID NOW TESTING** |  |  |  |  |
| Assessment centre | Symptomatic (n=70,879) | 34.7 (34.4, 0.04-101) | 40.5% | 49.9% |
|  | Asymptomatic** (n=10,818) | 34.5 (32.4, 0.1-102) | 46.8% | 40.7% |
| Hospital | Symptomatic (n=16,924) | 50.7 (53.6, 0.0007-107.9) | 48.0% | 90.2% |
| emergency shelters and addiction treatment facilities | Symptomatic (n=1,927) | 42.2 (39.2, 2.7-101.2) | 60.8% | 14.2% |
| Mobile unit | Symptomatic (n=4,224) | 31.8 (31.1, 0.001-103.9) | 38.9% | 15.5% |
|  | Asymptomatic (n=19,578) | 51.4 (47.8, 0.5-107.2) | 43.5% | 56.0% |
| **COMBINED OROPHARYNGEAL AND NASAL SWAB FOR ID NOW TESTING** |  |  |  |  |
| Assessment centre | Symptomatic (n=3890) | 35.6 (35.9, 0.7-98.3) | 40.8% | 68.7% |
|  | Asymptomatic** (n=681) | 31.2 (29.1, 3.4-83.1) | 48.6% | 76.8% |
| emergency shelters and addiction treatment facilities | Symptomatic (n=191) | 40.6 (39.4, 19.1-68.2) | 62.8% | 1.1% |

NP: nasopharyngeal
\*Either NP or oropharyngeal swab was used for parallel RT-PCR testing. In hospitalized patients, a minority of samples were other specimen types such as endotracheal tube aspirates.
\*\*Asymptomatic close contacts only

**Table 2:** Performance of ID NOW, compared to RT-PCR, using oropharyngeal swabs or combined oropharyngeal and nasal (O+N) swabs.

| Oropharyngeal Swab |  |  |  |
| --- | --- | --- | --- |
| Assessment Centre: Symptomatic |  |  |  |
|  |  | RT-PCR |  |
|  |  | Positive | Negative |
| ID | Positive | 9833 | 315 |
| NOW | Negative | 800 | 59,931 |
| Assessment Centre: Asymptomatic |  |  |  |
|  |  | RT-PCR |  |
|  |  | Positive | Negative |
| ID | Positive | 768 | 73 |
| NOW | Negative | 140 | 9,837 |
| Emergency Shelters and Addiction Treatment Facilities |  |  |  |
|  |  | RT-PCR |  |
|  |  | Positive | Negative |
| ID | Positive | 62 | 8 |
| NOW | Negative | 19 | 1838 |
| Hospital |  |  |  |
|  |  | RT-PCR |  |
|  |  | Positive | Negative |
| ID | Positive | 2,624 | 97 |
| NOW | Negative | 308 | 13,895 |
| Mobile: Symptomatic |  |  |  |
|  |  | RT-PCR |  |
|  |  | Positive | Negative |
| ID | Positive | 529 | 22 |
| NOW | Negative | 72 | 3,601 |
| Mobile: Asymptomatic |  |  |  |
|  |  | RT-PCR |  |
|  |  | Positive | Negative |
| ID | Positive | 365 | 78 |
| NOW | Negative | 129 | 19,006 |

| O+N Swab |  |  |  |
| --- | --- | --- | --- |
| Assessment Centre: Symptomatic |  |  |  |
|  |  | RT-PCR |  |
|  |  | Positive | Negative |
| ID | Positive | 335 | 18 |
| NOW | Negative | 28 | 3,509 |
| Assessment Centre: Asymptomatic |  |  |  |
|  |  | RT-PCR |  |
|  |  | Positive | Negative |
| ID | Positive | 35 | 5 |
| NOW | Negative | 10 | 631 |
| Emergency Shelters and Addiction Treatment Facilities |  |  |  |
|  |  | RT-PCR |  |
|  |  | Positive | Negative |
| ID | Positive | 3 | 0 |
| NOW | Negative | 1 | 187 |

**Figure 1:**
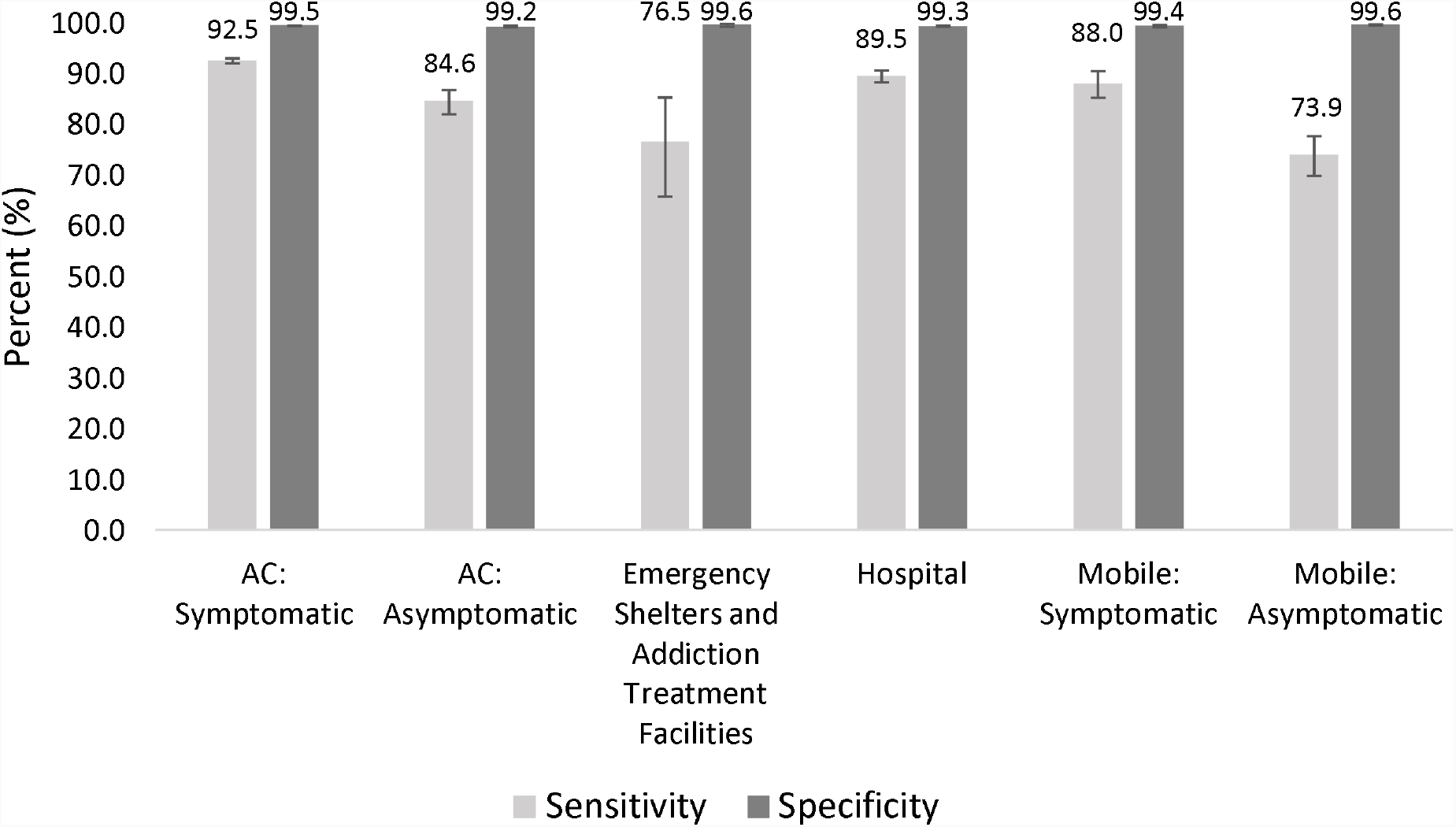
Sensitivity and specificity of ID NOW (oropharyngeal swab) compared to RT-PCR (oropharyngeal swab or nasopharyngeal swab). Error bars represent 95% confidence intervals. AC = assessment centre.

Positive predictive value (PPV) was highest for symptomatic individuals presenting to assessment centres (PPV 96.9%, 95% CI 96.5-97.2%), followed by symptomatic patients in hospital or emergency rooms (96.4%, 95% CI 95.7-97.1%), symptomatic patients tested via mobile units (96.0%, 94.1-97.3%), asymptomatic individuals presenting to assessment centres (91.3%, 95% CI 89.2-93.1%), symptomatic individuals at emergency shelters and addiction treatment facilities (88.6%, 95% CI 79.3-94.0%) and asymptomatic patients tested via mobile units (82.4%, 95% CI 78.8-85.5%). Negative predictive value (NPV) was highest for asymptomatic patients tested via mobile units (99.3%, 95% CI 98.8-99.1%), symptomatic individuals at emergency centres and addiction treatment facilities (99.0%, 95% CI 98.5-99.3%), symptomatic individuals presenting to assessment centres (98.7%, 95% CI 98.6-98.8%), asymptomatic individuals presenting to assessment centres (98.6%, 95% CI 98.3-98.8%), symptomatic patients tested via mobile units (98.0%, 95% CI 97.6-98.4%), and symptomatic patients in hospital or emergency rooms (97.8%, 95% CI 97.6-98.0%).

Ct values for ID NOW true positive and true negative results is provided in Table 3. Mean E gene Ct values were significantly higher among ID NOW false negatives compared to true positives and among asymptomatic compared to symptomatic populations, with the exception of mean E gene Ct value of false negatives between symptomatic vs asymptomatic in mobile testing and in assessment centres using O+N swabs (p>0.05).

**Table 3:**
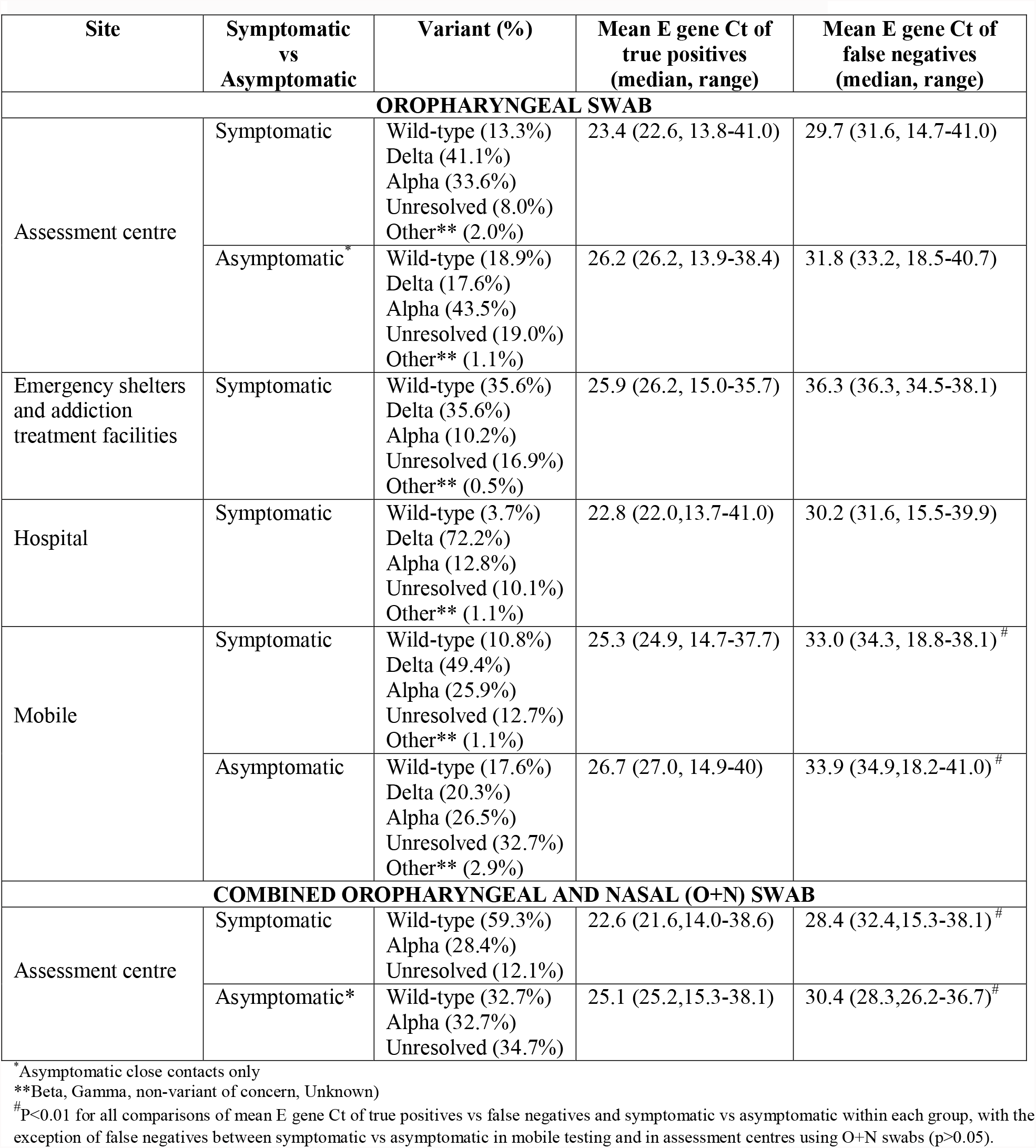
Cycle threshold of ID NOW true positives (ID NOW +, RT-PCR +) and false negatives (ID NOW -, RT-PCR +) between sites and populations tested.

| Site | Symptomatic<br>vs<br>Asymptomatic | Variant (%) | Mean E gene Ct of<br>true positives<br>(median, range) | Mean E gene Ct of<br>false negatives<br>(median, range) |
| --- | --- | --- | --- | --- |
| **OROPHARYNGEAL SWAB** |  |  |  |  |
| Assessment centre | Symptomatic | Wild-type (13.3%)<br>Delta (41.1%)<br>Alpha (33.6%)<br>Unresolved (8.0%)<br>Other** (2.0%) | 23.4 (22.6, 13.8-41.0) | 29.7 (31.6, 14.7-41.0) |
|  | Asymptomatic* | Wild-type (18.9%)<br>Delta (17.6%)<br>Alpha (43.5%)<br>Unresolved (19.0%)<br>Other** (1.1%) | 26.2 (26.2, 13.9-38.4) | 31.8 (33.2, 18.5-40.7) |
| Emergency shelters<br>and addiction<br>treatment facilities | Symptomatic | Wild-type (35.6%)<br>Delta (35.6%)<br>Alpha (10.2%)<br>Unresolved (16.9%)<br>Other** (0.5%) | 25.9 (26.2, 15.0-35.7) | 36.3 (36.3, 34.5-38.1) |
| Hospital | Symptomatic | Wild-type (3.7%)<br>Delta (72.2%)<br>Alpha (12.8%)<br>Unresolved (10.1%)<br>Other** (1.1%) | 22.8 (22.0,13.7-41.0) | 30.2 (31.6, 15.5-39.9) |
| Mobile | Symptomatic | Wild-type (10.8%)<br>Delta (49.4%)<br>Alpha (25.9%)<br>Unresolved (12.7%)<br>Other** (1.1%) | 25.3 (24.9, 14.7-37.7) | 33.0 (34.3, 18.8-38.1)<sup>#</sup> |
|  | Asymptomatic | Wild-type (17.6%)<br>Delta (20.3%)<br>Alpha (26.5%)<br>Unresolved (32.7%)<br>Other** (2.9%) | 26.7 (27.0, 14.9-40) | 33.9 (34.9,18.2-41.0)<sup>#</sup> |
| **COMBINED OROPHARYNGEAL AND NASAL (O+N) SWAB** |  |  |  |  |
| Assessment centre | Symptomatic | Wild-type (59.3%)<br>Alpha (28.4%)<br>Unresolved (12.1%) | 22.6 (21.6,14.0-38.6) | 28.4 (32.4,15.3-38.1)<sup>#</sup> |
|  | Asymptomatic* | Wild-type (32.7%)<br>Alpha (32.7%)<br>Unresolved (34.7%) | 25.1 (25.2,15.3-38.1) | 30.4 (28.3,26.2-36.7)<sup>#</sup> |

\*Asymptomatic close contacts only
\*\*Beta, Gamma, non-variant of concern, Unknown)
<sup>#</sup>P<0.01 for all comparisons of mean E gene Ct of true positives vs false negatives and symptomatic vs asymptomatic within each group, with the exception of false negatives between symptomatic vs asymptomatic in mobile testing and in assessment centres using O+N swabs (p>0.05).

There was no significant difference in sensitivity and specificity of ID NOW when comparing symptomatic and asymptomatic patients under 5 years of age, under 10 years of age, under 18 years of age, and over 18 years of age (Supplementary Table 1-4). There was no significant difference in sensitivity and specificity of ID NOW when comparing symptomatic and asymptomatic patients based on gender (Supplementary Table 5-6).

There was no significant difference in sensitivity when a NP or OP swab for confirmatory RT-PCR (or variant) testing was done among symptomatic or asymptomatic patients (Supplementary Table 7-8). However, there was slight differences in specificity, with OP swabs having higher specificity compared to NP swabs. There was no significant difference in sensitivity between symptomatic or asymptomatic patients with alpha compared to the delta variant (Supplementary Table 9-10).

### ID NOW testing using combined oropharyngeal and nasal swabs

4,762 paired samples were analyzed (412 RT-PCR positive). 4,571 samples were from 3 assessment centres and 191 samples were collected from 1 urban emergency shelter. Baseline characteristics of these samples is provided in Table 1. ID NOW results, compared to RT-PCR, are provided in Table 2. There was no statistically significant differences in sensitivity or specificity noted between results from ID NOW using oropharyngeal swabs and those using O+N swabs (Figure 2).

**Figure 2:**
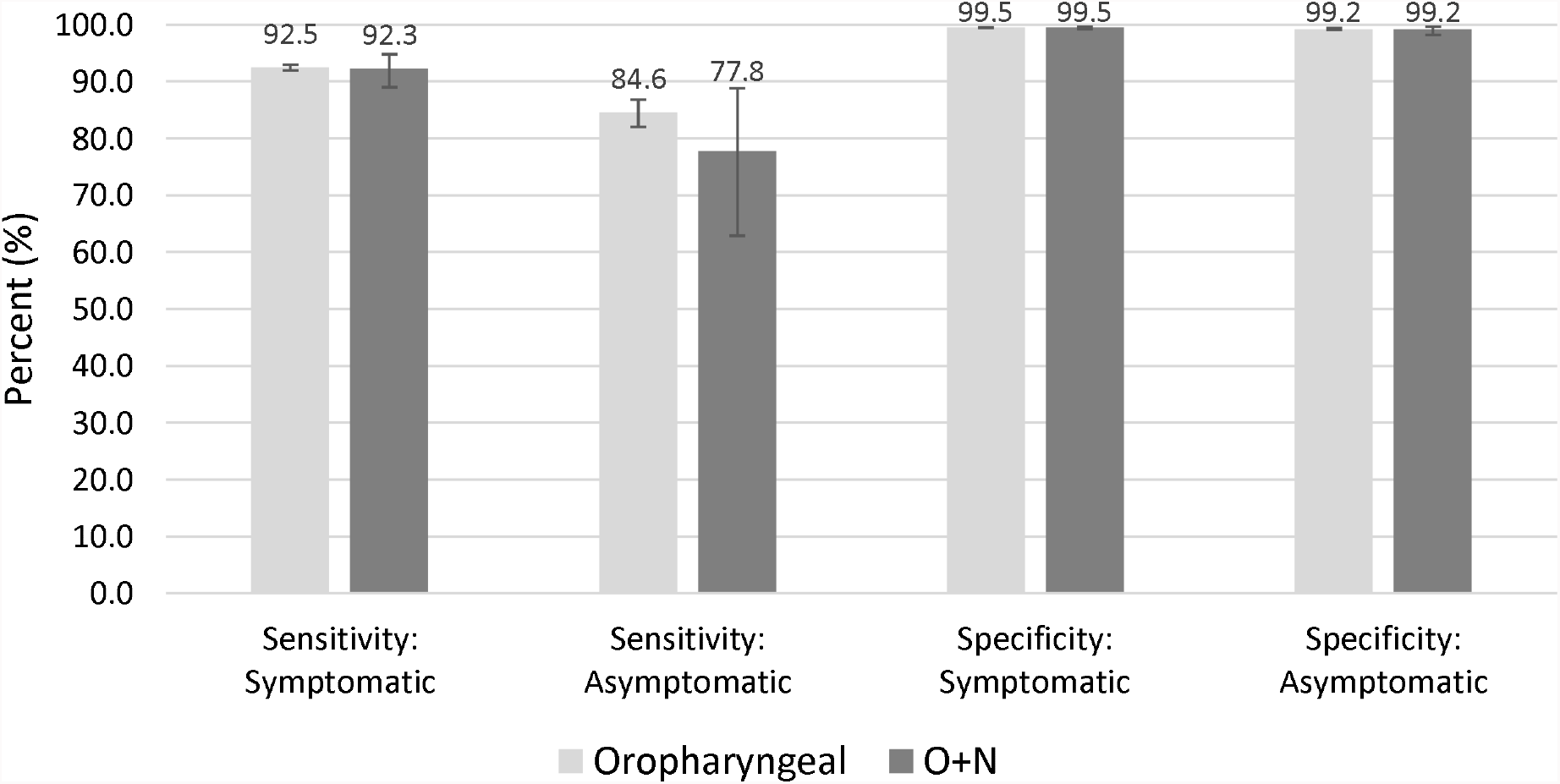
Sensitivity and specificity of ID NOW, compared to RT-PCR, for ID NOW specimens collected using either oropharyngeal swab or combined oropharyngeal and nasal (O+N) swab at COVID-19 assessment centres. Error bars represent 95% confidence intervals.

## DISCUSSION

Use of point of care SARS-CoV-2 tests, such as the ID NOW, for the detection of SARS-CoV-2 among individuals remains a worthwhile endeavour. Even if confirmatory testing of negatives is performed, identifying positives at the point of care has several advantages. It can speed important public health measures, such as contact tracing and isolation. Moreover, it has significant benefits for the laboratory in terms of decreasing contamination error and improving laboratory processes. For instance, decreasing the number of positive samples entering the laboratory can decrease the risk of false positive results by reducing the probability of SARS-CoV-2 contamination during RT-PCR testing. In addition, the decrease in positive samples can improve efficiencies in other laboratory processes, such as pooling.

In our large, multicentre, population-based study, we observed differing ID NOW sensitivities, compared to RT-PCR, based on the population being tested. Sensitivity was highest (92.5%) when tested among symptomatic individuals presenting to community COVID-19 assessment centres and lowest for asymptomatic individuals associated with community outbreaks (73.9%, 95% CI 69.8-77.7%, n=494 RT-PCR positive). These results highlight that the ID NOW performance is not standard across populations tested, likely as a result of its poorer performance with lower viral loads, as can be observed among asymptomatic individuals or individuals who are further out from their symptom onset.^5^ This outcome can better help direct where ID NOW devices are most suitable, and provide information to consider when goals of testing are reviewed. As expected, we observed lower average E gene Ct values among symptomatic individuals compared to asymptomatic individuals and among ID NOW false negative compared to true positive samples. Higher Ct values observed in results from emergency shelters and addiction treatment facilities is likely a manifestation of the unique population being tested. While testing was limited to individuals with symptom onset within the first 7 days, it can be challenging to differentiate those with symptoms in these settings, particularly those who spend frequent time outside in the cold (i.e. rhinorrhea), and have fluctuating symptoms related to other factors such as drug use.

ID NOW sensitivity was slightly lower in the symptomatic populations tested using a mobile service or in-hospital based settings. There may be various factors to account for this. Firstly, ID NOW testing is performed immediately after sample collection at assessment centres, whereas hospitals and mobile service often require transportation to the on-site lab or mobile unit prior to ID NOW testing. While ID NOW testing was mandated to be done within 1 hour from collection, short periods of time from transportation may potentially affect performance.^10^ Secondly, a higher proportion of individuals with lower viral loads may be tested in hospitals and via mobile. For mobile testing (mostly outbreaks), individuals with symptoms were possibly early in their symptom onset at time of testing, whereas those presenting to hospitals were possibly late in their symptom onset; both of which can be associated with higher Ct values and, therefore, lower viral loads.^5^ Symptomatic individuals presenting to assessment centres, in comparison, are often within the first few days of symptom onset, but because it generally takes approximately 24 hours to arrange a booking, are not at the very beginning of their symptom onset.

There are clear differences in ID NOW sensitivity with respect to symptoms, with asymptomatic populations having an approximately 10% decrease in sensitivity. There were significantly higher mean E gene Ct values observed when testing asymptomatic versus symptomatic individuals in assessment centres. This has important implications when using the ID NOW for screening of asymptomatic populations, where the implications of a rapid test for such a population needs to be carefully considered and weighed over the risks of a substantial decrease in sensitivity compared to other testing methods (RT-PCR). However, the sensitivity of the ID NOW compared to other point of care options, including rapid antigen tests, is far superior, with the sensitivity of rapid antigen tests ranging from 30-50% in asymptomatic populations.^4^

ID NOW sensitivity in our study is underestimated due to the large number (1,822, 11.3% of positives) of ID NOW positives that were excluded from our analysis, 93% of which came from symptomatic individuals from assessment centres. These were excluded due to the lack of parallel RT-PCR testing. If these excluded samples were included as true positives, sensitivity of symptomatic individuals presenting to assessment centres would increase to 93.5% (95% CI 93.0-93.9%).

ID NOW specificity was consistently high across the populations tested (>99%), and is consistent with prior studies within the literature.^4^ When comparing ID NOW to RT-PCR from individuals with confirmed COVID-19, we previously demonstrated a significant number of ID NOW positive, RT-PCR negative discrepant results.^10^ This suggests that other factors may account for discrepant results, such as sampling error. It is noted that multiple individuals within our dataset had an ID NOW positive result followed by subsequent negative RT-PCR, but then on repeat RT-PCR the next day were positive. These patients were considered ID NOW false positives in our study, although in reality would be considered true positives based on this information. As such, ID NOW specificity is likely higher than what we have reported. Although the exclusion of many ID NOW positive results in our study may contribute to overestimation of ID NOW specificity, this would be primarily limited to symptomatic individuals at assessment centres, and would have little to no impact on the specificity observed among other populations studied.

Using an oropharyngeal swab or O+N swab for ID NOW testing yielded similar results among both symptomatic and asymptomatic populations. Given the higher inconvenience with O+N swabs, and the lack of advantages demonstrated over typical specimens, we recommend against using O+N swabs for ID NOW COVID-19 testing.

Strengths of this study include the large sample size and including both symptomatic and asymptomatic populations. We also studied testing in various real world locations, including community COVID-19 assessment centres, hospitals (inpatient and ED), emergency shelters and addiction treatment facilities, and continuing care and industry outbreaks. Due to the introduction of mandatory testing of many positive ID NOW or RT-PCR samples for variants of concern, which included E gene RT-PCR from a consistent RT-PCR testing platform (APL LDT), we were able to assess the specificity of the ID NOW and examine the relationship of E gene Ct values between true positive and false negative ID NOW results.

Our study had several limitations. Due to the heterogeneity of our populations tested, it is difficult to exclude confounders that may have contributed to ID NOW performance. However, no differences were observed in sensitivity based on common patient, collecting and testing characteristics, including age, gender, swab used, and variants of concern detected. Specificity was increased for RT-PCR samples collected using oropharyngeal swabs, but this difference was slight and limited to symptomatic populations. Exclusion of some ID NOW positive results from our study because of no parallel RT-PCR testing did have an impact on ID NOW performance, as evidenced by improved sensitivity (93.5% vs 92.5%), among symptomatic community patients presenting to assessment centres, when including these positive samples. Reasons behind missing parallel RT-PCR are multifactorial and include sample lost or discarded prior to testing, testing sites going against guidelines and not obtaining a second swab for RT-PCR confirmation, and patient demographic mismatches resulting in test cancellation or inability to match ID NOW and RT-PCR tests together in our electronic database. These excluded samples, however, would have little impact on ID NOW sensitivity and specificity outside of symptomatic individuals presenting to assessment centres. For instance, no significant changes in sensitivity among asymptomatic community patients that presented to assessment centres were observed when the 1822 excluded positive results were subsequently analyzed.

In conclusion, the performance of the ID NOW differs across populations tested for SARS-CoV-2. ID NOW performance can be maximized if testing is limited to symptomatic individuals presenting to community testing sites (assessment centres). Sensitivity drops by approximately 10% when the testing population is asymptomatic. ID NOW performance is not affected by age, gender, or variants of concern (alpha, delta). ID NOW testing with combined oropharyngeal + nasal swab had no effect on performance.

## Supporting information

Publication

## Data Availability

All data produced in the present study are available upon reasonable request to the authors

## Data Availability

All data produced in the present study are available upon reasonable request to the authors

## Acknowledgments

This work was funded using internal operating funds of Alberta Precision Laboratories and Alberta Health Services. Test kits and instruments were paid for by the Public Health Agency of Canada. We thank staff at AHS assessment centre, mobile teams, and staff at shelters and addiction recovery sites for collecting and testing samples in the community, and Alberta Precision Laboratory staff for assistance with testing of samples, and in the development and support of the various testing programs.

## Declarations

### Availability of data

Available upon request.

