## Supplementary material for "Then and NOW: A Prospective Population-Level Validation of the Abbott ID NOW SARS-CoV-2 Device Implemented in Multiple Settings for Testing Asymptomatic and Symptomatic Individuals": Publication

**Performing an Oropharyngeal+Nasal (O+N) Swab**

A) Remove any mucous from the patient's nose, with a tissue or cotton tipped swab prior to collecting the oropharyngeal and nasal swab.
B) Open the ID NOW swab for O+N collection.
C) Collect an oropharyngeal specimen using the ID NOW swab, as per standard protocol
D) Remove the ID NOW swab from the oropharynx and immediately use this swab to collect a nasal swab as per below.
E) Insert the ID NOW swab into one nostril of the patient. The swab tip should be inserted up to 2.5 cm (1 inch) from the edge of the nostril.
F) Roll the swab 3 times along the mucosa inside the nostril to ensure that both mucus and cells are collected.
G) Using the same ID NOW swab, repeat this process for the other nostril to ensure that an adequate sample is collected from both nasal cavities.
H) Remove the swab and place it into the ID NOW instrument for rapid testing.

Supplementary Table 1: Performance of ID NOW, compared to RT-PCR, using oropharyngeal swabs on outpatient children under age 5 years. Not all ID NOW positive samples had RT-PCR samples performed.

| | | | |
|---|---|---|---|
| **Symptomatic** | | | |
| | | RT-PCR | |
| | | Positive | Negative |
| ID NOW | Positive | 185 | 13 |
| | Negative | 13 | 2,435 |
| Sensitivity: 89.4% (95% CI 84.4-93.2%)<br>Specificity: 99.5% (95% CI 99.1-99.7%)<br>PPV: 93.4% (95% CI 89.2-96.1%)<br>NPV: 99.1% (95% CI 98.2-99.1%) | | | |
| **Asymptomatic** | | | |
| | | RT-PCR | |
| | | Positive | Negative |
| ID NOW | Positive | 36 | 5 |
| | Negative | 9 | 380 |
| Sensitivity: 80.0% (95% CI 65.4-90.4%)<br>Specificity: 98.7% (95% CI 97.0-99.6%)<br>PPV: 87.8% (95% CI 74.9-94.6%)<br>NPV: 97.7% (95% CI 95.9-98.7%) | | | |

CI: Confidence interval

Supplementary Table 2: Performance of ID NOW, compared to RT-PCR, using oropharyngeal swabs on outpatient children under age 10 years. Not all ID NOW positive samples had RT-PCR samples performed.

| **Symptomatic** | | | |
|---|---|---|---|
| | | RT-PCR | |
| | | Positive | Negative |
| ID NOW | Positive | 590 | 35 |
| | Negative | 64 | 6,796 |
| Sensitivity: 90.2% (95% CI 87.7-92.4%)<br>Specificity: 99.5% (95% CI 99.3-99.6%)<br>PPV: 94.4% (95% CI 92.4-95.9%)<br>NPV: 99.1% (95% CI 98.8-99.3%) | | | |
| **Asymptomatic** | | | |
| | | RT-PCR | |
| | | Positive | Negative |
| ID NOW | Positive | 120 | 10 |
| | Negative | 34 | 1,403 |
| Sensitivity: 77.9% (95% CI 70.5-84.2%)<br>Specificity: 99.3% (95% CI 98.7-99.7%)<br>PPV: 92.3% (95% CI 86.6-95.7%)<br>NPV: 97.6% (95% CI 96.8-98.2%) | | | |

CI: Confidence interval

Supplementary Table 3: Performance of ID NOW, compared to RT-PCR, using oropharyngeal swabs on outpatient children under age 18 years. Not all ID NOW positive samples had RT-PCR samples performed.

| **Symptomatic** | | | |
|---|---|---|---|
| | | RT-PCR | |
| | | Positive | Negative |
| ID NOW | Positive | 1,807 | 69 |
| | Negative | 174 | 13,925 |
| Sensitivity: 91.2% (95% CI 89.9-92.4%)<br>Specificity: 99.5% (95% CI 99.4-99.6%)<br>PPV: 96.3% (95% CI 95.4-97.1%)<br>NPV: 98.8% (95% CI 98.3-98.7%) | | | |
| **Asymptomatic** | | | |
| | | RT-PCR | |
| | | Positive | Negative |
| ID NOW | Positive | 309 | 37 |
| | Negative | 91 | 4,406 |
| Sensitivity: 77.3% (95% CI 72.8-81.3%)<br>Specificity: 99.2% (95% CI 98.9-99.4%)<br>PPV: 89.3% (95% CI 85.8-92.0%)<br>NPV: 98.0% (95% CI 97.6-98.3%) | | | |

CI: Confidence interval

Supplementary Table 4: Performance of ID NOW, compared to RT-PCR, using oropharyngeal swabs on outpatient adults over age 18 years. Not all ID NOW positive samples had RT-PCR samples performed.

| **Symptomatic** | | | |
|---|---|---|---|
| | | RT-PCR | |
| | | Positive | Negative |
| ID NOW | Positive | 8,555 | 268 |
| | Negative | 698 | 49,605 |
| Sensitivity: 92.5% (95% CI 91.9-93.0%)<br>Specificity: 99.5% (95% CI 99.4-99.5%)<br>PPV: 97.0% (95% CI 96.6-97.3%)<br>NPV: 98.6% (95% CI 98.3-98.5%) | | | |
| **Asymptomatic** | | | |
| | | RT-PCR | |
| | | Positive | Negative |
| ID NOW | Positive | 824 | 114 |
| | Negative | 178 | 24,434 |
| Sensitivity: 82.2% (95% CI 79.7-84.6%)<br>Specificity: 99.5% (95% CI 99.4-99.6%)<br>PPV: 87.9% (95% CI 85.7-89.7%)<br>NPV: 99.3% (95% CI 99.2-99.4%) | | | |

CI: Confidence interval

Supplementary Table 5: Performance of ID NOW, compared to RT-PCR, using oropharyngeal swabs on outpatient males.

| **Symptomatic** | | | |
|---|---|---|---|
| | | RT-PCR | |
| | | Positive | Negative |
| ID NOW | Positive | 4,825 | 121 |
| | Negative | 386 | 24,900 |
| Sensitivity: 92.6% (95% CI 91.9-93.3%)<br>Specificity: 99.5% (95% CI 99.4-99.6%)<br>PPV: 97.6% (95% CI 97.1-97.9%)<br>NPV: 98.5% (95% CI 98.3-98.6%) | | | |
| **Asymptomatic** | | | |
| | | RT-PCR | |
| | | Positive | Negative |
| ID NOW | Positive | 607 | 72 |
| | Negative | 135 | 12,596 |
| Sensitivity: 81.8% (95% CI 78.8-84.5%)<br>Specificity: 99.4% (95% CI 99.3-99.6%)<br>PPV: 89.4% (95% CI 87.0-91.4%)<br>NPV: 98.9% (95% CI 98.8-99.1%) | | | |

CI: Confidence interval

Supplementary Table 6: Performance of ID NOW, compared to RT-PCR, using oropharyngeal swabs on outpatient females.

| Symptomatic | | | |
|---|---|---|---|
| | | RT-PCR | |
| | | Positive | Negative |
| ID NOW | Positive | 5,485 | 216 |
| | Negative | 479 | 38,404 |
| Sensitivity: 92.0% (95% CI 91.3-92.7%)<br>Specificity: 99.4% 95% CI (99.4-99.5%)<br>PPV: 96.2% (95% CI 95.7-96.7%)<br>NPV: 98.8% (95% CI 98.3-98.6%) | | | |
| **Asymptomatic** | | | |
| | | RT-PCR | |
| | | Positive | Negative |
| ID NOW | Positive | 510 | 79 |
| | Negative | 130 | 15,868 |
| Sensitivity: 79.7% (95% CI 76.4-82.7%)<br>Specificity: 99.5% (95% CI 99.4-99.6%)<br>PPV: 86.6% (95% CI 83.8-89.0%)<br>NPV: 99.2% (95% CI 98.6-98.9%) | | | |

CI: Confidence interval

Supplementary Table 7: Performance of ID NOW, compared to RT-PCR, when nasopharyngeal swabs are used for RT-PCR for outpatients.

| Assessment Centre: Symptomatic | | | |
|---|---|---|---|
| | | RT-PCR | |
| | | Positive | Negative |
| ID NOW | Positive | 4,222 | 191 |
| | Negative | 431 | 29,175 |
| Sensitivity: 90.7% (95% CI 89.9-91.6%)<br>Specificity: 99.4% (95% CI 99.3-99.4%)<br>PPV: 95.7% (95% CI 95.1-96.2%)<br>NPV: 98.5% (95% CI 98.4-98.7%) | | | |
| **Assessment Centre: Asymptomatic** | | | |
| | | RT-PCR | |
| | | Positive | Negative |
| ID NOW | Positive | 313 | 50 |
| | Negative | 128 | 14,531 |
| Sensitivity: 71.0% (95% CI 66.5-75.2%)<br>Specificity: 99.7% (95% CI 99.6-99.8%)<br>PPV: 86.2% (95% CI 82.5-89.3%)<br>NPV: 99.1% (95% CI 99.0-99.2%) | | | |

CI: Confidence interval

Supplementary Table 8: Performance of ID NOW, compared to RT-PCR, when oropharyngeal swabs are used for RT-PCR for outpatients.

| **Assessment Centre: Symptomatic** | | | |
|---|---|---|---|
| | | RT-PCR | |
| | | Positive | Negative |
| ID NOW | Positive | 4,574 | 82 |
| | Negative | 404 | 31,965 |
| Sensitivity: 91.9% (95% CI 91.1-92.6%)<br>Specificity: 99.7% (95% CI 99.7-99.8%)<br>PPV: 98.2% (95% CI 97.8-98.6%)<br>NPV: 98.8% (95% CI 98.6-98.8%) | | | |
| **Assessment Centre: Asymptomatic** | | | |
| | | RT-PCR | |
| | | Positive | Negative |
| ID NOW | Positive | 495 | 35 |
| | Negative | 135 | 14,139 |
| Sensitivity: 78.6% (95% CI 75.2-81.7%)<br>Specificity: 99.8% (95% CI 99.7-99.8%)<br>PPV: 93.4% (95% CI 91.0-95.2%)<br>NPV: 99.1% (95% CI 98.9-99.2%) | | | |

CI: Confidence interval

Supplementary Table 9: Performance of ID NOW, compared to RT-PCR, for outpatients with the delta variant.

| **Symptomatic** | | | |
|---|---|---|---|
| | | RT-PCR | |
| | | Positive | Negative |
| ID NOW | Positive | 4,569 | N/A |
| | Negative | 142 | N/A |
| Sensitivity: 97.0% (95% CI 96.5-97.5%) | | | |
| **Asymptomatic** | | | |
| | | RT-PCR | |
| | | Positive | Negative |
| ID NOW | Positive | 263 | N/A |
| | Negative | 15 | N/A |
| Sensitivity: 94.6% (95% CI 91.3-97.0%) | | | |

CI: Confidence interval

Supplementary Table 10: Performance of ID NOW, compared to RT-PCR, for outpatients with the alpha variant.

| **Symptomatic** | | | |
|---|---|---|---|
| | | RT-PCR | |
| | | Positive | Negative |
| ID NOW | Positive | 3,633 | N/A |
| | Negative | 130 | N/A |
| Sensitivity: 96.6% (95% CI 95.9-97.1%) | | | |
| **Asymptomatic** | | | |
| | | RT-PCR | |
| | | Positive | Negative |
| ID NOW | Positive | 505 | N/A |
| | Negative | 52 | N/A |
| Sensitivity: 90.7% (95% CI 87.9-93.0%) | | | |

CI: Confidence interval
